# A Phase 2 open-label study to determine the safety and efficacy of weekly dosing of ATL1102 in patients with non-ambulatory Duchenne muscular dystrophy

**DOI:** 10.1101/2022.01.16.22269029

**Authors:** IR Woodcock, G Tachas, N Desem, PJ Houweling, EM Yiu, M Kean, J Emmanuel, R Kennedy, K Carroll, K de Valle, J Adams, SR Lamandé, C Coles, C Tiong, D Villano, P Button, J-Y Hogrel, S Catling-Seyffer, MB Delatycki, MM Ryan, ATL1102 in DMD clinical trial group

## Abstract

**Background:** ATL1102 is a 2’MOE gapmer antisense oligonucleotide to the CD49d alpha subunit of VLA-4. ATL1102 inhibits expression of CD49d on lymphocytes, thereby reducing their survival, activation and migration to sites of inflammation. Children with Duchenne muscular dystrophy (DMD) have dystrophin deficient muscles. These are susceptible to contraction induced injury which triggers the immune system, exacerbating muscle damage. CD49d is a biomarker of disease severity in DMD, with increased numbers of high CD49d expressing T cells correlating with more severe and progressive weakess, despite corticosteroid treatment.

**Methods:** This Phase 2 open label study assessed the safety, efficacy and pharmacokinetic profile of ATL1102 administered as 25 mg weekly by subcutaneous injection for 24 weeks in 9 non-ambulatory boys with DMD aged 10–18 years. Participants receiving corticosteroid therapy were allowed into the study if on a stable dose for at least 3 months. The main objective was to assess safety and tolerability of ATL1102. Secondary objectives included the effect of ATL1102 on lymphocyte numbers in the blood, functional changes in upper limb function as assessed by Performance of Upper Limb test (PUL 2.0) and upper limb strength using MyoGrip and MyoPinch compared to baseline.

**Results:** ATL1102 was generally safe and well tolerated. No serious adverse events were reported. There were no participant withdrawals from the study. The most commonly reported adverse events were injection site erythema and skin discoloration. There was no statistically significant change in lymphocyte count from baseline to week eight (mean change -0.56×10^9^/L 95%CI -1.52, 0.40), week twelve (mean change -0.53×10^9^/L 95%CI -1.65, 0.58) or week twenty-four (mean change -0.28×10^9^/L 95%CI -1.10, 0.55) of dosing however, the CD3+CD49d+ T lymphocytes were statistically significantly higher at week 28 compared to week 24, four weeks past the last dose (mean change 0.40×10^9^/L 95%CI 0.05, 0.74; p=0.030). Functional muscle strength, as measured by the PUL2.0, EK2 and Myoset grip and pinch measures, and MRI fat fraction of the forearm muscles were stable throughout the trial period.

**Conclusion:** ATL1102, a novel antisense drug being developed for the treatment of inflammation that exacerbates muscle fibre damage in DMD, appears to be safe and well tolerated in non-ambulant boys with DMD. The apparent stabilisation observed on multiple muscle disease progression parameters assessed over the study duration support the continued development of ATL1102 for the treatment of DMD.

## Introduction

Duchenne muscular dystrophy (DMD), a severe, progressive, X-linked genetic muscle disease is the most common muscle disorder in boys, affecting 1 in 5000 live male births worldwide.(1) Boys with DMD have onset of progressive muscle weakness in the first decade of life, with death due to cardiorespiratory failure expected in the late third or early fourth decades.(2) Currently the only disease modifying medical treatment is corticosteroid therapy, which delays loss of ambulation by a median 3 years, to 13 years of age(3-5) but carries a significant treatment burden of adverse effects.(5)

DMD is associated with absence of dystrophin from muscle. This causes increased susceptibility to contraction-induced muscle damage, with activation of the innate immune macrophages in turn activating the adaptive immune system T lymphocytes, leading to upregulation of pro-inflammatory cytokines, including the extracellular structural protein osteopontin, resulting in chronic inflammation, fibrosis and reduced muscle strength.(6) CD49d, the alpha chain subunit of integrin very late antigen 4 (VLA-4) is expressed widely on immune cells in this cascade and can bind osteopontin. (7) In patients with DMD, the number of CD49d high expressing T lymphocytes is inversely proportional to ambulation speed, with highest concentration seen in non-ambulant patients.(8) Patients with higher concentrations have more severe weakness and are more likely to lose ambulation before 10 yrs of age despite corticosteroid use, suggesting CD49d may be a biomarker of disease severity or activity.(9) In ex vivo studies a monoclonal antibody to VLA-4 prevented patient T-cell binding to muscle cells and transendothelial migration, highlighting a potential therapeutic avenue.(9)

ATL1102 is a second-generation immunomodulatory 2’MOE gapmer antisense oligonucleotide which specifically targets human CD49d RNA. After binding to the RNA of CD49d, intracellular RNase H attaches resulting in downregulation of CD49d RNA. ATL1102 has previously been trialled to treat Relapsing Remitting Multiple Sclerosis (RRMS), noting that CD49d high expressing T cells are the effector and central memory T cells in RRMS. In this phase 2 RRMS clinical trial, ATL1102 dosed 200mg three times weekly in the first week and twice weekly to 8 weeks substantially reduced inflammatory brain lesions by 88.5% and circulating lymphocytes and T lymphocytes by 25%.(10)

Reported here, collaborators from the same institution ran two separate but complementary studies. The initial trial, a phase 2 clinical trial examining for the first time the safety and efficacy of a low dose ATL1102 treatment for 24 weeks in non-ambulant patients with DMD on concomitant corticosteroid treatment. The second, a pre-clinical study in the *mdx* mouse model for DMD, conducted using a mouse specific second generation CD49d ASO (ISIS 348574) to show that monotherapy treatment can reduce CD49d mRNA expression in muscle and decrease contraction induced muscle damage.

## Methods

### Ethics statement

The clinical trial received approval from the Royal Children’s Hospital Human Research Ethics Committee with assigned number HREC/17/RCHM/121. The trial was subsequently registered at the Australian New Zealand Clinical Trials Registry (ACTRN12618000970246). An independent Data Safety Monitoring Board (DSMB) was established to provide safety oversight for the trial. Pre-clinical *mdx* mouse analyses were approved by the Murdoch Children’s Research Institute (MCRI) animal care and ethics committee (ACEC; approval number A899).

### Pre-clinical studies in mice

The *mdx* mouse model is commonly used to study DMD. We tested efficacy of the second generation 2’MOE gapmer mouse specific CD49d ASO ISIS 348574 as ATL1102 is specific to human CD49d RNA and not homologous to mouse. Mdx mice do not have circulating lymphocytes with high CD49d but have high CD49d expressing lymphocytes in the lymph nodes at 9 weeks.(11) Symptomatic 9 week old *mdx* mice were treated for 6 weeks to determine the effect of ISIS 348574 on *CD49d* mRNA expression and muscle function measures.

Male *mdx* (n = 48) and age matched C57Bl10/J wild-type (WT, n = 12) controls were purchased from JAX laboratories at 5 weeks of age and acclimatised to the MCRI facility for a total of 4 weeks. Animals were housed in a specific-pathogen-free environment at a constant ambient temperature of 22°C and 50% humidity on a 12 h light-dark cycle, with *ad libitum* access to food and water. The *mdx* mice were randomly assigned to 4 treatment groups (saline, low (5mg/kg) and high (20mg/kg) dose ISIS 348574, and a control gapmer oligonucleotide with the same 20 nucleotides scrambled such that it is not complementary to CD49d or other RNA (20mg/kg)). Mice (n = 12 /group) received weekly subcutaneous injections of saline, ISIS348574 and control oligonucleotide for a total of 6 weeks. The high dose (20mg/kg/week) equates to 1.6mg/kg/w dose in human equivalent body surface area (BSA) and the low dose (5mg/kg/week) equates to 0.4mg/kg/week dose on BSA. After treatment mice were anaesthetised using inhaled isoflurane (0.6 ml per min) and muscle function was examined using the Arora Scientific 1300A whole mouse test system and 701C stimulator as previously published.(12) Mice were then euthanised by cervical dislocation and the spleen and skeletal muscle (quadriceps) were collected for further analysis.

### Flow cytometry

The *mdx* spleen was perforated to isolate the splenic sub-cellular content and incubated in red blood cell lysis buffer (Thermofisher) for ten minutes at 4°C. Splenocytes were centrifuged 1500 g for five minutes at 4°C. Cell pellets were washed in wash buffer (PBS:1% BSA (bovine albumin serum)). CD4+ and CD8+ T cell populations were identified using anti-CD4-V450 (Biolegend, San Diego, CA, USA) and anti-CD8a-allophycocyanin – cyanine 7 (anti-CD8a-APC-Cy7) (Biolegend, San Diego, CA, USA). Stained cells were analysed using BD LSRFortessa™ X-20 Cell Analyzer to identify populations of CD4+ and CD8+ T cells.

### Clinical Trial Design

This was a phase 2 open-label clinical trial assessing the safety of ATL1102 in non-ambulant boys with DMD concomitantly with standard of care corticosteroid therapy. Inclusion criteria for the clinical trial are included in the supplementary data Figure 1.

Participants received weekly subcutaneous injections of 25mg of ATL1102 for twenty-four weeks. Injections were administered into subcutaneous fat of the abdomen by a registered nurse or trained parent. Injection sites were rotated in quadrants around the umbilicus. Parents monitored injection sites for cutaneous reactions and participant discomfort for fourty-eight hours post-administration. Adverse events were recorded in a diary which was returned to the study coordinators at each fortnightly visit.

Participants underwent fortnightly venepuncture for exploratory and safety blood tests. This included monitoring of haematology, biochemistry and inflammatory markers and at point of care urinalysis dipstick to monitor kidney function. Participants were seen monthly by the study team for physical examination and respiratory function assessments.

Participant safety was monitored routinely and at regularly scheduled meetings by the independent DSMB.

### Outcome Measurements

The primary endpoint of the trial was safety of ATL1102 as assessed by the frequency and intensity of adverse events, including injection site reactions and any laboratory value derangement.

Secondary outcome measures included both laboratory and functional efficacy endpoints. Laboratory efficacy outcome measures included lymphocyte-modulation activity determined by cell surface flow cytometry measuring variation in the number and percentage of total lymphocytes as well as those CD4 and CD8 T lymphocytes expressing high levels of CD49d (CD49dhi) to week twenty-four of treatment and to week twenty-eight, four weeks past the last treatment.

Changes in upper limb muscle strength and function were measured at baseline and again at weeks five, eight, twelve and twenty-four. Muscle function was assessed by a questionnaire-based outcome measure of disease burden (Egen Klassifikation Scale version 2 - EK2) and performance-based measures (the Performance of the Upper Limb scale version 2 (PUL 2.0), and the Moviplate 30 second finger tapping score of the MyoSet tool). The Myoset tool also measured distal upper limb strength as determined by the MyoPinch (key pinch strength) and MyoGrip (hand grip strength) scores.(13-15) Other outcome measures assessed were respiratory function (forced vital capacity (FVC) and forced expiratory volume in one second (FEV1)), and quality of life assessed using the neuromuscular module of the Pediatric Quality of Life Instrument (PedsQL NMD™).

### Muscle Magnetic Resonance Imaging (MRI)

Participants underwent MRI of the dominant forearm at baseline, week twelve and week twenty-four. Unilateral upper-limb MRI was performed at 1.5T (Siemens Aera; Siemens, Erlangen, Germany) using a flexible surface matrix coil (4-Channel Flex Coil) wrapped around the forearm. Participants lay in the scanner in the head-first supine position, with the arm to be imaged lying in a comfortable position on the scanner bed alongside the torso. Two point-Dixon images were acquired (3D gradient-echo TE1/TE2/TR = 2.39/4.44/6.99ms, flip angle 10°, nine 6mm axial slices, slice gap 0mm, FOV 18×18cm, matrix 320×320, pixel size 0.56×0.56mm, NEX = 4). Fat fraction maps were obtained using on scanner tools.

Change over time in muscle composition (atrophy, oedema and fatty infiltration) was measured on the muscles of the central, proximal, and distal forearm using Short Tau Inversion Recovery (STIR) and 3-point Dixon sequences on MRI. Changes in muscle composition were scored using the semi-quantitative visual scoring Mercuri method and by quantitative fat fraction analysis.(16-18) Due to fatty infiltration, identification of individual muscles was challenging, such that a compartment composite score of volar, dorsal and ECRLB Br (extensor carpi radialis longus/brevis and brachioradialis) compartments was used as per a previous published study.(19) The lean muscle mass was calculated using previously published methods: Cross-sectional muscle compartment area x ((100 – total muscle compartment fat percent) / 100).(19)

### Statistical analysis

Based upon data from a previous study of ATL1102 in RRMS patients analyzing blood 3 days after the last dose in week 8, the laboratory efficacy end point of lymphocyte modulation potential was established as a reduction in total lymphocyte count of 0.47×10^9^/L (25% reduction).(10) For the sample size calculation, the level of significance was set to 0.05 with a 2-sided paired t-test, mean difference of 0.47 (×10^9^/L) from baseline to end of treatment, and standard deviation of 0.428 (×10^9^/L). A sample size of 9 participants was calculated as required to achieve a power of 80%. Data were analysed using SAS® Version 9.4. primary and secondary efficacy measures were analysed using the paired t-test and the non-parametric Wilcoxon sign-rank test. The study was not powered to see a change on the secondary efficacy endpoints from baseline to end of treatment.

## Results

### Proof of concept preclinical study using mdx mice

*Ex vivo* analyses of monocytes collected from *mdx* mice (n = 3), showed that the ASO to mouse CD49d, ISIS 348574 can reduce the expression of CD49d mRNA (Fig. 1A). We then examined the *in vivo* response to ISIS 348574 in *mdx* mice treated for 6 weeks which showed that *CD49d* mRNA expression was reduced in skeletal muscle by approximately 40% when treated with either a low (5mg/kg/week) or high (20mg/kg/week) dose of ISIS 348574, compared to saline controls (Fig. 1B, One-way ANOVA with Tukey correction, p = * <0.05, ** <0.01).

**Figure 1:**
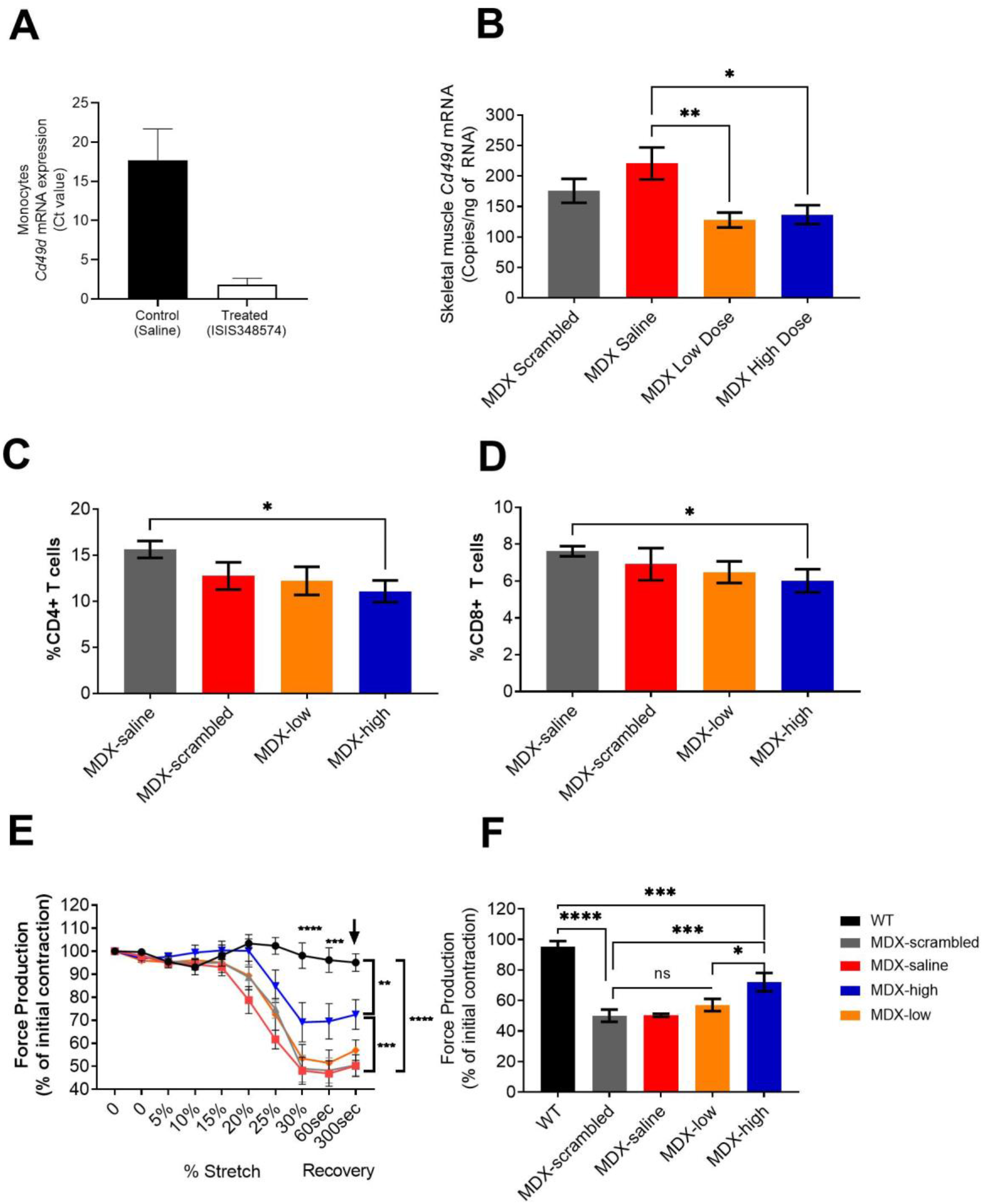
Pre-clinical data using the mdx mouse model of DMD to test the effects of ISIS 348574 (mouse specific Cd49d oligonucleotide to ATL 1102) in vivo. A) Monocytes were isolated from the spleens of *mdx* mice (n = 3) and exposed to a single dose of ISIS 348574 for 48hrs *in vitro* to show that we could achieve a reduction in *CD49d* mRNA using ISIS 348574 to mouse CD49d RNA. B) Following 6 weeks of treatment *CD49d* mRNA expression was reduced in mice treated with either the low (5mg/kg/week) or high (20mg/kg/week) dose of ISIS 348574, compared to saline controls (One-way ANOVA with Tukey correction, p = * <0.05, ** <0.01). C and D) Proportion of CD4+ and CD8+ T cells from the spleens of *mdx* mice with and without ISIS 348574 drug treatment. Cells are expressed as a proportion of total live cells isolated from the spleen. One way ANOVA with Fishers LSD test, * p < 0.05. E and F) *In situ* muscle physiology analyses shows that *mdx* mice treated with either saline (red, ∼45% force recovery), scrambled (grey, ∼45% force recovery) or low dose (orange, ∼ 50% force recovery) ISIS 348574 were susceptible to eccentric muscle contraction damage compared to wild-type (black) controls, whereas the mice treated with a high dose of ISIS 348574 were resistant to the effects of eccentric muscle damage and produced 72% of the original force following the eccentric muscle damaging protocol. This was still significantly less than the 95% force recovery seen in WT mice, however this improvement in force following a muscle damage protocol suggests that the use of a 20mg/kg/week dose of ISIS 348574 was able to protect the muscles of *mdx* mice. One-way ANOVA with Fishers LSD test, p = * <0.05, ** <0.01, *** <0.001), ****<0.0001).

This study also found that *mdx* mice treated with the 20mg/kg/week dose of ISIS 348574 showed a reduction in the percentage of splenic CD4+ (30%, p<0.05) and CD8+ (21%, p<0.05) T lymphocytes, compared to saline treated mice (Fig. 1C and D). Furthermore *in situ* muscle funcation analyses show that the high dose (20mg/kg/week) treated *mdx* mice were protected from the effects of eccentric muscle damage, producing 72% of the original muscle force (SEM <u>+6,</u> P<0.01). This was in contrast to *mdx* mice treated with either saline, scrambled control or low dose (5mg/kg) ISIS 348574, which generated approximately 50% of the original force following eccentric muscle contractions. (Fig 1. E and F, One-way ANOVA with Fishers LSD test, p = * <0.05, ** <0.01, *** <0.001).

### Clinical Trial Results

Eleven adolescent males with DMD were screened for participation. All had been non-ambulant for at least six months prior to screening. Two screened participants were excluded (one participant had started cardioprotective medication within three months of the initial visit and the other participant exceeded the pre-determined weight limit for inclusion).

Nine participants were enrolled into the open-label study. All had a confirmed pathogenic variant in DMD, with a clinical phenotype consistent with DMD as assessed by their treating/referring clinician and the study investigator. Participant demographics are summarized in Table 1.

**Table 1:** Summary of Participant Demographics

| Characteristic | Category | Statistic | ATL1102<br>N = 9 |
| --- | --- | --- | --- |
| Sex | Male | n (%) | 9 (100) |
| Age (years) |  | Mean (SD) | 14.9 (2.1) |
|  |  | Median (range) | 14.0 (12 - 18) |
| Weight (kg) |  | Mean (SD) | 52.7 (9.8) |
| Height (cm) |  | Mean (SD) | 141.1 (10.0) |
| BMI |  | Mean (SD) | 27.1 (7.4) |
| Time since non-ambulant (years) |  | Median (range) | 2.2 (0.6 – 9.2) |
| Corticosteroid Medication | Yes | n (%) | 8 (88.9) |
|  | *Prednisolone* |  | 3 (33.3) |
|  | *Deflazacort* |  | 5 (55.6) |

### Safety

There were no serious adverse events (SAEs) or suspected or unexpected serious adverse reactions (SUSARs). A total of 136 adverse events were recorded (table 2), with all participants reporting at least one adverse event. Sixty-three percent of the reported adverse events were injection site related, with all but one participant experiencing transient erythema within twenty-four hours of the administration of ATL1102. Six (67%) participants had mild post-inflammatory hyperpigmentation of the skin of their abdomen which was persistent; four had resolved and two were ongoing but improving at the post completion follow-up study visit. The hyperpigmentation was noticed in the first participant after receiving eleven weekly doses of ATL1102. The DSMB was made aware and the participant informed consent form updated. In the five subsequent participants who had a similar reaction it was seen after four to eleven doses. The hyperpigmentation was not regarded as a clinical safety concern by the DSMB. Pain, discomfort or atrophy of the subcutaneous tissues were not reported, and there were no signs of systemic involvement. No participants withdrew from the study. There were no other significant adverse events felt to be related to ATL1102 or its administration.

**Table 2:**
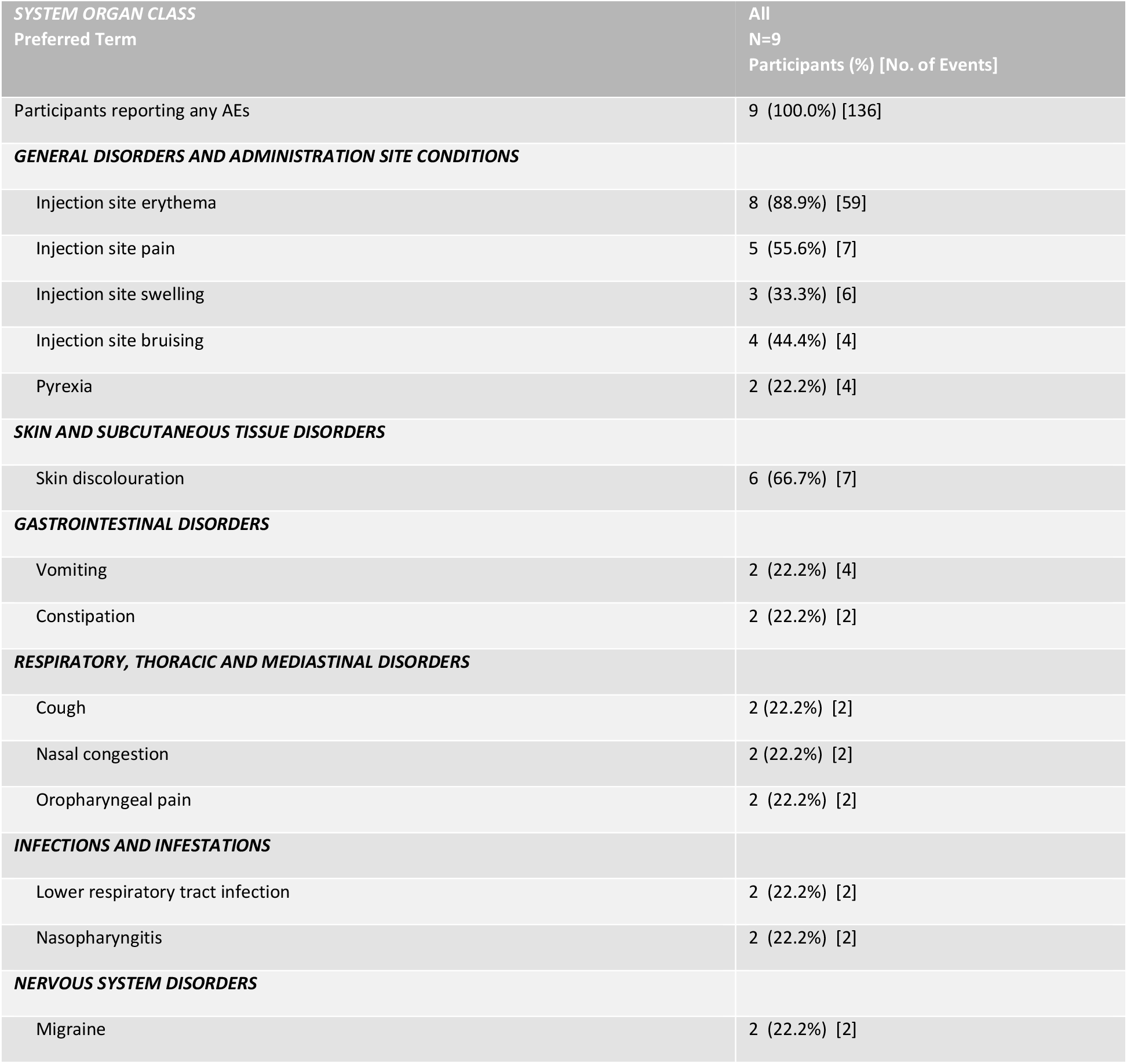
Treatment Emergent Adverse Events reported in at least two participants

| **SYSTEM ORGAN CLASS**<br>**Preferred Term** | **All**<br>**N=9**<br>**Participants (%) [No. of Events]** |
| --- | --- |
| Participants reporting any AEs | 9 (100.0%) [136] |
| **GENERAL DISORDERS AND ADMINISTRATION SITE CONDITIONS** |  |
| Injection site erythema | 8 (88.9%) [59] |
| Injection site pain | 5 (55.6%) [7] |
| Injection site swelling | 3 (33.3%) [6] |
| Injection site bruising | 4 (44.4%) [4] |
| Pyrexia | 2 (22.2%) [4] |
| **SKIN AND SUBCUTANEOUS TISSUE DISORDERS** |  |
| Skin discolouration | 6 (66.7%) [7] |
| **GASTROINTESTINAL DISORDERS** |  |
| Vomiting | 2 (22.2%) [4] |
| Constipation | 2 (22.2%) [2] |
| **RESPIRATORY, THORACIC AND MEDIASTINAL DISORDERS** |  |
| Cough | 2 (22.2%) [2] |
| Nasal congestion | 2 (22.2%) [2] |
| Oropharyngeal pain | 2 (22.2%) [2] |
| **INFECTIONS AND INFESTATIONS** |  |
| Lower respiratory tract infection | 2 (22.2%) [2] |
| Nasopharyngitis | 2 (22.2%) [2] |
| **NERVOUS SYSTEM DISORDERS** |  |
| Migraine | 2 (22.2%) [2] |

### Efficacy

#### Lymphocyte Count

There was no statistically significant decrease in lymphocyte count from baseline to week eight, week twelve or week twenty-four of dosing (Table 3). There was no statistically significant decrease in CD49d+CD3+CD8^+^ or CD49d+CD3+CD4^+^ T lymphocytes seen between baseline, weeks 8, 12 or 24 (table 3). This 9 participant trial did not achieve the pre-specified laboratory activity outcome measure of a significant -0.47×10^9^/L (25% reduction) in total lymphocyte count.

**Table 3:**
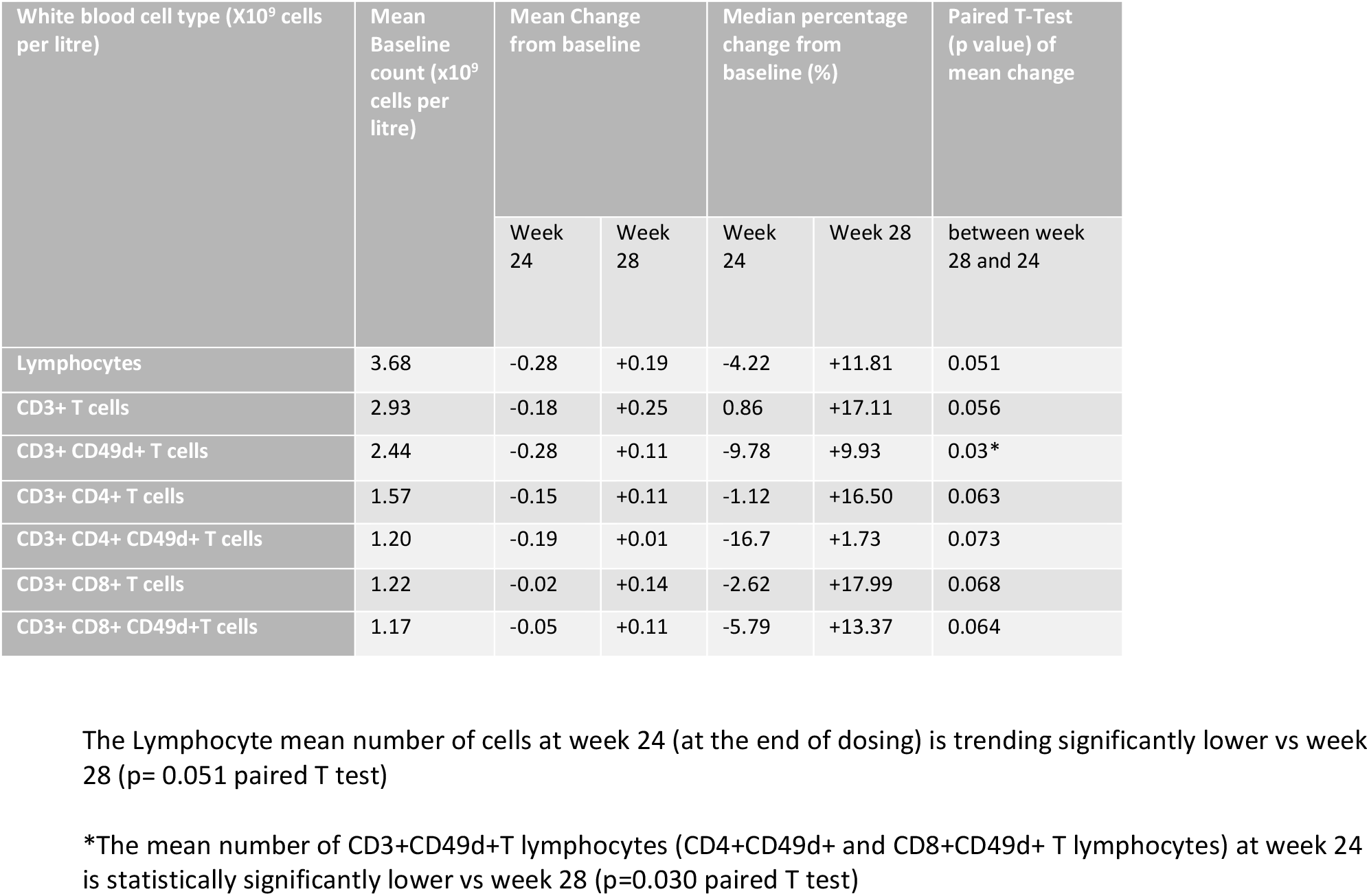
Summary of lymphocytes mean change from baseline to weeks 24 and 28

There was, though, a consistent trend toward declines in the mean number of lymphocytes, and CD49d+ T lymphocytes measured 3 days post-dose at week 8, 12 and 24. The mean number of CD3+CD49d+ T lymphocytes (i.e. CD3+CD4+ and CD3+CD8+ expressing CD49d) measured at week 28 was statistically significantly higher compared to end of dosing at week 24 (mean change 0.40×10^9^/L 95%CI 0.05, 0.74; paired T-Test, p=0.030) (Table 3). Repeated measures analysis of CD3-CD49d+ NK lymphocytes in a *post hoc* analysis comparing baseline to a linear combination of values measured three days post-dose at weeks 8, 12 and 24 was significantly lower compared to baseline (p=0.018), with comparable NK lymphocyte numbers at week 28.

#### Functional Outcome Measures

There were no statistically significant changes in any upper limb functional outcome measures at week 24 compared to baseline (table 4). The PUL2.0 score remained stable with no significant change between baseline and week twenty-four with a mean increase in PUL2.0 score (less disease impact) of 0.9 (95%CI -1.33, 3.11). EK2 scores were stable throughout the trial period. ATL1102 treatment showed no significant effect on lung function throughout the 24 week trial period (table 4). The components of the Myoset all reflected stable grip and pinch strength over the course of the trial.

**Table 4:** Change in Functional Outcome Measures from baseline to week 24.

| Change from Baseline to Week 24 |  |  |  |  |  |  |  |  |  |
| --- | --- | --- | --- | --- | --- | --- | --- | --- | --- |
| Patient No. | PUL 2.0 | MyoGrip (dom) (Kg) | MyoGrip (dom) (% Pred) | MyoPinch (dom) (Kg) | MyoPinch (dom) (% Pred) | MoviPlate Score (dom) | % Predicted FVC | % Predicted PEF | EK2* |
| 01-001 | +2 | -0.63 | -4.49 | 0.03 | -0.62 | -14.0 | -3.20 | 6.30 | +1 |
| 01-002 | +2 | 0.22 | 0.49 | -0.02 | -0.29 | 13.0 | -14.8 | -17.3 | +1 |
| 01-003 | 0 | 0.68 | 1.02 | -0.40 | -6.59 | -3.0 | -9.10 | 8.70 | +2 |
| 01-004 | +2 | 1.09 | 1.01 | 0.37 | 2.99 | 7.0 | 0.80 | 7.20 | +2 |
| 01-006 | -3 | -0.27 | -0.60 | 0.07 | 0.94 | 8.0 | -6.50 | 6.90 | -6 |
| 01-008 | +7 | 1.00 | 1.11 | 0.30 | 2.77 | 3.0 | -7.70 | -18.2 | -1 |
| 01-009 | 0 | -0.33 | -3.75 | -0.22 | -4.97 | 7.0 | -9.10 | -4.30 | +2 |
| 01-010 | 0 | 0.05 | 0.11 | 0.06 | 0.72 | -15.0 | -0.40 | 9.20 | -1 |
| 01-011 | -2 | 0.11 | -1.31 | -0.18 | -3.63 | 11.0 | -1.10 | 2.00 | +2 |
| Mean Change (95% CI): | 0.9 (-1.33, 3.11) | 0.2 (-0.25, 0.67) | -0.7 (-2.33, 0.90) | 0.0 (-0.18, 0.19) | -1.0 (-3.56, 1.63) | 1.9 (-6.08, 9.85) | -5.68 (-9.60, -1.76) | 0.06 (-8.33, 8.44) | 0.2 (-1.80, 2.25) |
| *Higher score = greater disability |  |  |  |  |  |  |  |  |  |
| #Reduction in Fat Fraction (%) = improvement |  |  |  |  |  |  |  |  |  |

#### MRI

In this trial, there was some variation in the quality of proximal and distal slices due to variable positioning of the participant’s forearm in consecutive scans. The quality of the central slices through the muscle body in the forearms muscles was not compromised and so this measurement was chosen for detailed comparison between baseline and week 24 scans for each participant. No significant change was apparent between baseline and week twenty-four for the mean Mercuri semi-quantitative score of fatty infiltration (0.1 point change), atrophy (0 point change) and muscle oedema (0.3 point change) measuring the central slice around the elbow (table 6). There was a trend towards minor improvement in the percentage fat fraction in all muscle groups measuring the central slice, although statistical significance was not achieved. There was no significant pattern of change in cross-sectional muscle area of any muscle group (table 5).

**Table 5:** Change in the MRI Central reading fat fraction, cross sectional area and lean muscle mass from baseline to week 24.

| Mean Change (95% CI) from Screening/Baseline to Week 24 |  |  |
| --- | --- | --- |
| MRI Parameter | N | MRI Central Reading Mean (95%CI) |
| **Fat Fraction (%)** |  |  |
| Volar Muscle | 9 | -0.57 (-7.81, 6.68) |
| Dorsal Muscles | 9 | -0.88 (-3.41, 1.65) |
| ECRLB-Br | 9 | -0.12 (-6.42, 6.17) |
| Average Fat Fraction | 9 | -0.52 (-5.62, 4.58) |
| **Cross Sectional Muscle Area (mm<sup>2</sup>)** |  |  |
| Volar Muscle | 9 | 22.78 (-31.2, 76.73) |
| Dorsal Muscles | 9 | 0.89 (-18.9, 20.65) |
| ECRLB-Br | 9 | -1.33 (-8.94, 6.28) |
| Total Area | 9 | 22.33 (-36.8, 81.42) |
| Lean Muscle Mass (mm<sup>2</sup>) | 9 | 13.9 (72.6, 100.4) |

ECRLB-Br = extensor carpi radialis longus/brevis and brachioradialis.
**Volar Muscles:** flexor digitorum profundus and flexor pollicis longus (FDP), flexor digitorum superficialis and palmaris longus (FDS), flexor carpi ulnaris (FCU), flexor carpi radialis (FCR)
**Dorsal Muscles:** Extensor carpi ulnaris (ECU), extensor digiti minimi (EDM), extensor digitorum (ED), extensor pollicis longus (EPL), abductor pollicis longus (APL), extensor carpi radialis longus/brevis and brachioradialis (ECRLB-Br), but the ECRLB-Br are not included in the Dorsal muscle measurement in the Central Reading
Change in the MRI Proximal reading average Fat Fraction (%) from baseline to week 24 was -2.14 [95%CI -7.60; 3.3] for the 9 patients.

**Table 6:** Mercuri visual semi-quantative score in whole forearm compartment from baseline to week 24.

| Whole Forearm |  |  |  |
| --- | --- | --- | --- |
|  | Baseline | Wk 24 | Change |
| Fatty Infiltration | 5.2 | 5.3 | 0.1 |
| Atrophy | 1.6 | 1.6 | 0.0 |
| Oedema | 2.6 | 2.9 | 0.3 |

#### Correlation of parameters assessed in the Phase 2 study

Correlation analyses were performed across assessment measures PUL2.0, Myoset and MRI. Positive correlations were observed in the Phase 2 study between the different measures of muscle function of Moviplate scores and the PUL 2.0 scores of the distal domain (*r* = 0.664) which support the consistency of the observed changes across the measures assessed in the study over the 24 week ATL1102 treatment period (See supplementary data Figure 2).

Positive correlations were also observed in the Phase 2 study between the MRI results of the lean muscle area (non-fat) and MyoGrip results (*r* = 0.604), suggesting a consistency of results across the different parameters of muscle structure and muscle strength (See supplementary data Figure 3).

## Discussion

### Safety

This open-label phase 2 clinical trial met its primary safety end point. All but one participant experienced post-injection site erythema, swelling or discomfort suggesting that the investigation product is a mild irritant, as has been observed with other MOE antisense drugs, and is commonly reported with subcutaneiously injected drugs. Future clinical trials of ATL1102 could consider including using ice as a pre-injection site treatment to minimise these reactions. Six participants experienced an unexpected post-inflammatory skin hyperpigmentation which resolved or faded at the completion of the study. Interestingly this reaction has not been previously reported in other clinical trials of ATL1102 and was not viewed as a safety concern.(10)

The dose chosen for this 24 week trial (25mg/week) was considered as a presumed safe dose in this patient population. In a previous phase 2 trial in individuals with RRMS a loading dose of 200mg every other day for one week was administered then 400mg/week (twice weekly 200mg) for seven weeks.(10) This DMD trial was the first clinical trial to investigate the safety of ATL1102 over a six-month period.

### Lymphocyte Modulation

Given the previously observed action of ATL1102 of reducing lymphocytes in the RRMS study, the trial sample size was calculated to see a 25% reduction in total lymphocyte count when the drug is at equilibrium from week 8. This activity endpoint was not met in this trial. There was however a consistent trend toward declines in the mean number of lymphocytes at week 8, 12 and 24 each measured 3 days past dosing, and statistically significant reductions in the mean number of CD49d+ NK lymphocytes at week 8, 12 and 24 weeks of treatment, using repeated measures analysis. The mean number of CD49d+ T lymphocytes (i.e CD3+CD4+ and CD3+CD8+ that are CD49d+) was statistically significantly higher at week 28 compared to week 24, indicating a rebound elevation of CD49d+ T lymphocytes four weeks post the last treatment dose. These results collectively suggest that ATL1102 suppresses CD49d expressing lymphocytes at a dose of 25mg per week. It is anticipated that higher doses will increase the level of lymphocyte reduction whilst maintaining a favourable safety profile in part due to sparing of the majority of T lymphocytes and NK lymphocytes. Future studies will look at dose escalation as supported by this study, and modelling with ATL1102.

### PUL2.0 and EK2 upper limb function

PUL2.0 measures shoulder, elbow, and wrist finger dimensions of disease burden and is a reliable measure of disease severity and progression in DMD. The mean change from baseline to week twenty-four in PUL2.0 was an increase in function by 0.9 (95% CI -1.33 to 3.11). Although the Minimal Clinical Important Difference (MCID) for the PUL2.0 has not been established, previously published data from a historical cohort suggested that over a twelve month period a mean decrease in PUL2.0 score of 2.17 can occur, albeit in a cohort not directly comparable to the participants in the phase 2 study due to older age and larger proportion not on corticosteroids.(20) An external historical cohort with same inclusion criteria as in the ATL1102 phase 2 trial, showed a decrease in PUL2.0 score of 2.0 (standard deviation 3.02) from baseline over a six month period.(21) In the ATL1102 phase 2 study four of the nine patients achieved an increase in their PUL2.0 score of +2, and another three patients were stabilized in the PUL2.0 score (Table 5). This is an encouraging trend that warrants further investigation.

There was no change in the EK2 over the course of the trial period. This composite outcome measure encompasses multiple aspects of disease burden and as such is a useful clinical monitoring tool (higher score equals greater disease burden) but is not likely to be as responsive as the PUL2.0 measure to small changes in upper limb function. As such, a stabilisation over the six month trial periods is encouraging and needs to be confirmed in future studies.

### Myoset Tests: MoviPlate, MyoGrip, and MyoPinch

The MyoSet functional outcome measures consist of the MoviPlate muscle function assessment of repetitive flexion extension of the wrist and fingers, and MyoGrip and MyoPinch assessment of muscle grip and pinch strength. (13, 14) These measures have been validated for use in clinical trials of DMD; MyoGrip and MyoPinch in particular have been shown to be sensitive to change in non-ambulant boys and to correlate well with lean muscle mass on MRI.(15) There was no change in any of these measures over the trial period. Previous natural history studies have shown significant deterioration over six months (Grip -0.5kg [95%CI -1.01; 0.002] and Pinch -0.38kg [95%CI -0.53; -0.22]). (18, (15) Matching the MyoGrip and MyoPinch protocol with that of a previously published natural history cohort allowed for comparison of change in grip and pinch strength over a six month period, yielding a statistically significant improvement on grip (p=0.03) and pinch (p=0.003) strength.(19) The lack of decline in these measures with ATL1102 during the trial period is once again encouraging and warrants further investigation.

### MRI of Upper Limb

Magnetic Resonance Imaging of muscle is increasingly used as a biomarker for disease stage and progression. The most widely used scoring method is the Mercuri Score, which requires a skilful investigator to visually score the chosen muscles based upon a standardised set of criteria encompassing degree of atrophy, oedematous changes and fatty infiltration of the muscle, to create an aggregate score. The more recent development of automated fat fraction analysis reduces the inter-user variability and provides a more quantitative measure of assessment. Matching the MRI protocol with that of a previously published natural history cohort allowed for direct comparison of change in fat fraction over a six month period.(19) From this published natural history data, disease is expected to progress with a mean increase of central forearm muscle fat fraction percentage of 3.9% (95%CI 1.9,5.7) over six months. The apparent trend towards a decrease in mean forearm muscle fat fraction of 0.52% (95% CI -5.62, 4.58; Median 1.4%) seen after six months of treatment with ATL1102 may suggest that ATL1102 could be modifying the rate of fatty infiltration into these muscles. Comparison of the mean change in fat fraction analysis of the central slice of the total muscle compartment between this study population and those of the natural history cohort published does not yield a significant difference (p=0.079). For future MRI studies it would be important to set a clearer protocol with imaging tags placed over surface landmarks to ensure uniformity of subsequent scans.

## Conclusion

The proof of concept pre-clinical data supports a potential protective effect of an antisense oligonucleotide to CD49d RNA in the mdx mouse model of DMD. This phase 2 open-label clinical trial has shown that ATL1102 has a good safety profile and is well tolerated with minor injection site reactions the only treatment-related adverse events reported. The positive observations in functional efficacy outcomes suggesting stabilization, and results compared with historical natural history data, particularly the PUL2.0, MyoGrip, MyoPinch and MRI fat fraction analysis, justifies the ongoing drug development program of ATL1102 in non-ambulant boys with DMD and provides a rationale to proceed with larger placebo-controlled studies of this novel therapeutic agent.

## Data Availability

All data produced in the present study are available upon request to the authors

## Acknowledgements

The authors wish to acknowledge the contribution of Isabelle Ledoux and Simone Birnbaum of the Insitute of Myology, Paris, France who provided support with the quality control and analysis of the MyoSet functional outcome measures; Valeria Ricotti of the Dubowitz Neuromuscular Centre, London UK, who provided support on the comparative analysis of the MRI observations; and Annabell Leske and Vicky Beal and the team at Avance Clinical, Adelaide, Australia.

## Supplementary Data

**Table 7:** Participant specific genetic variant within Dystrophin Gene

| Participant Number | Specific Genetic Variant within Dystrophin Gene |
| --- | --- |
| 1001 | Deletion of exons 48-50 |
| 1002 | splice site mutation in intron 45 |
| 1003 | deletion of exon 50 |
| 1004 | duplication within exon 15 causing premature stop codon |
| 1006 | deletion of exon 50 |
| 1008 | deletion of exons 8-27 |
| 1009 | nonsense substitution mutation in exon 23 causing premature stop codon |
| 1010 | nonsense mutation in exon 52 causing a premature stop codon |
| 1011 | deletion of exons 8-25 |

### Supplementary Data

Figure S1: Inclusion and Exclusion criteria for the clinical trial

Inclusion Criteria: Participants who:

1. Are adolescent males, aged 10 to 18 years inclusive, at the time of providing informed consent.
2. Have been diagnosed with Duchenne Muscular Dystrophy and have been non-ambulatory for at least 3 months. Non-ambulatory for this study is defined as having consistently required a wheelchair to mobilise more than a few metres for at least 3 months.
3. The diagnosis of DMD is to be confirmed by at least one of the following:
  a. Dystrophin immunofluorescence and/or immunoblot showing near complete dystrophin protein deficiency, and clinical picture consistent with typical DMD; or
  b. Gene deletion test positive (missing one or more exons) of the dystrophin gene, where the reading frame can be predicted as ‘out-offrame’, and clinical picture consistent with typical DMD; or
  c. Complete dystrophin gene sequencing showing an alteration (point mutation, duplication, or other mutation resulting in a stop codon) that can be definitely associated with DMD, with a typical clinical picture of DMD; or
  d. Positive family history of DMD confirmed by one of the criteria listed above in a sibling or maternal uncle, and clinical picture typical of DMD.
4. Have a body weight of more than 25 kg and less than or equal to 65 kg.
5. If currently receiving glucocorticoid therapy, have been on a stable dose of glucocorticoid therapy for at least 3 months prior to Day 1.
6. Are currently on stable doses of cardiac therapy (including angiotensin converting enzyme inhibitors, aldosterone receptor antagonists and/or beta blockers) for at least 3 months prior to Day 1.
7. Have a parent/guardian who is capable of understanding the purposes and risks of the study and able to provide written informed consent. If the participant is of sufficient maturity and has the ability to understand the nature and consequence of the study, involvement in study consent discussions are required.
8. Are able, and have a parent/guardian who are, willing and able to comply with scheduled visits, study drug administration plan, and study procedures.

Exclusion Criteria: Participants who:

1. Have been diagnosed with Duchenne Muscular Dystrophy and are still ambulatory. Ambulatory for this study is being able to complete at least 75 meters during the 6-minute walk test in the 4 weeks prior to Day 1.
2. Have the following abnormal haematology values during the Screening period or on Day 1, prior to first dose:
  a. Lymphocytes <1.2 × 109/L
  b. Neutrophils <1.8 × 109/L
  c. Platelets <150 × 109/L
3. Have a history of clinically significant bleeding or coagulation abnormalities.
4. Have hepatic dysfunction indicated by an abnormal total bilirubin and gamma glutamyl transferase (GGT) results at Screening.
5. Have renal impairment indicated by serum creatinine ≥ 1.5 mg/dL (132 umol/l) at Screening.
6. Have uncontrolled clinical symptoms and signs of congestive heart failure consistent with Stage C or Stage D criteria according to the American College of Cardiology/American Heart Association guidelines for cardiac dysfunction within 3 months of Day 1.
7. Have an inability to complete the cardiac, pulmonary or strength range of motion and mobility assessments at Screening.
8. Have taken nutritional, herbal, or antioxidant supplements that have a known demonstrated activity for maintaining or improving skeletal muscle strength or functional mobility within 4 weeks of Day 1. NOTE: daily multivitamin, Vitamin D or calcium supplements are permitted.
9. Are currently receiving antiplatelet or anticoagulant therapy, or have taken medication with an antiplatelet or anticoagulant effect within 4 weeks prior Day 1 (e.g., aspirin).
10. Have received any investigational product in the 2 months prior to Screening (4 months if the previous drug was a new chemical entity), whichever is longer.
11. Have severe behavioural disorder or inadequate cognitive development that would make them unable to comply with the study assessments, which, in the opinion of the investigator, makes the participant unsuitable for participation in the study.

**Figure S2:**
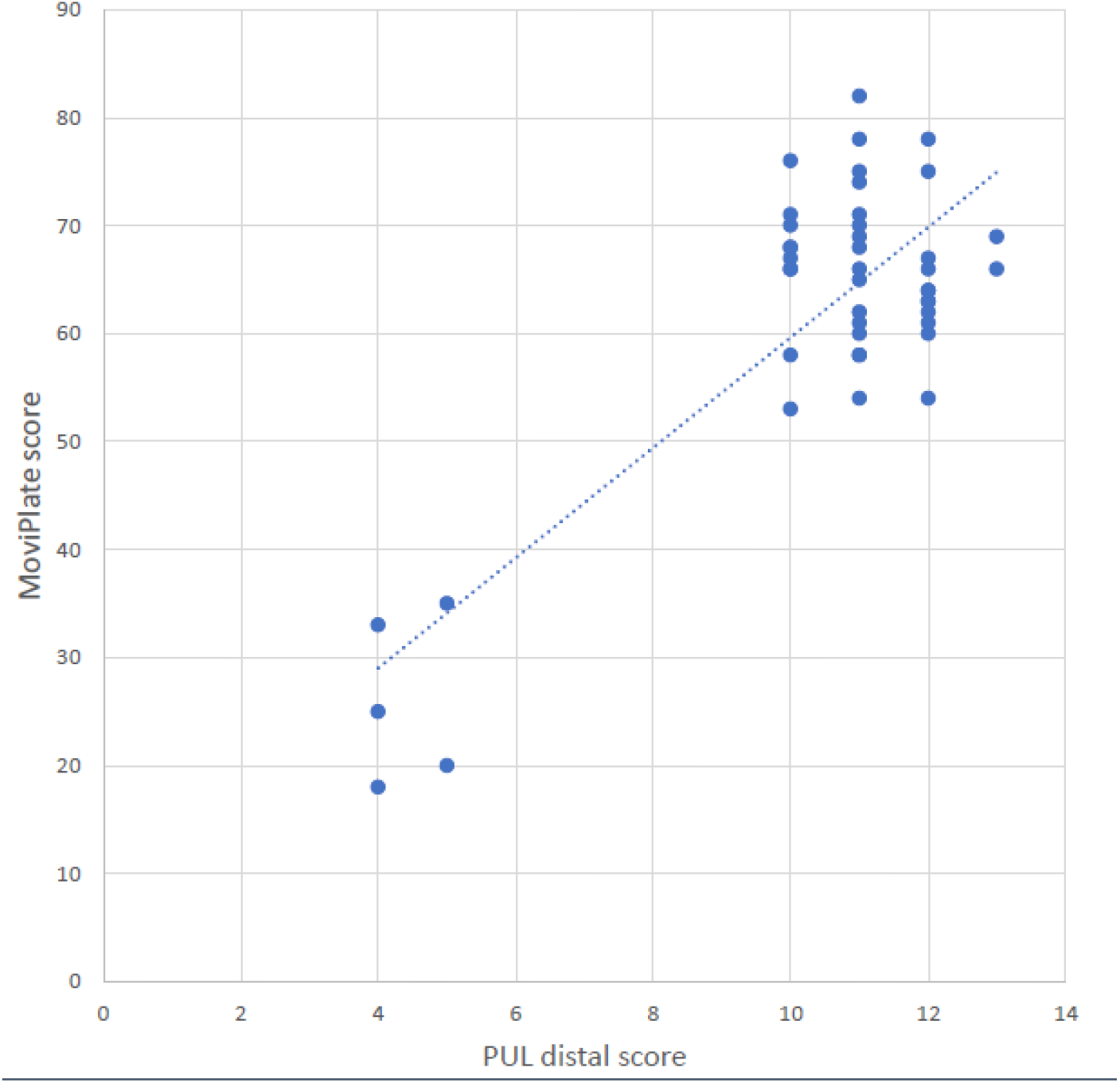
Correlation of the Moviplate scores and the PUL 2.0 distal dimension scores over the 24 Week ATL1102 treatment period:

**Figure S3:**
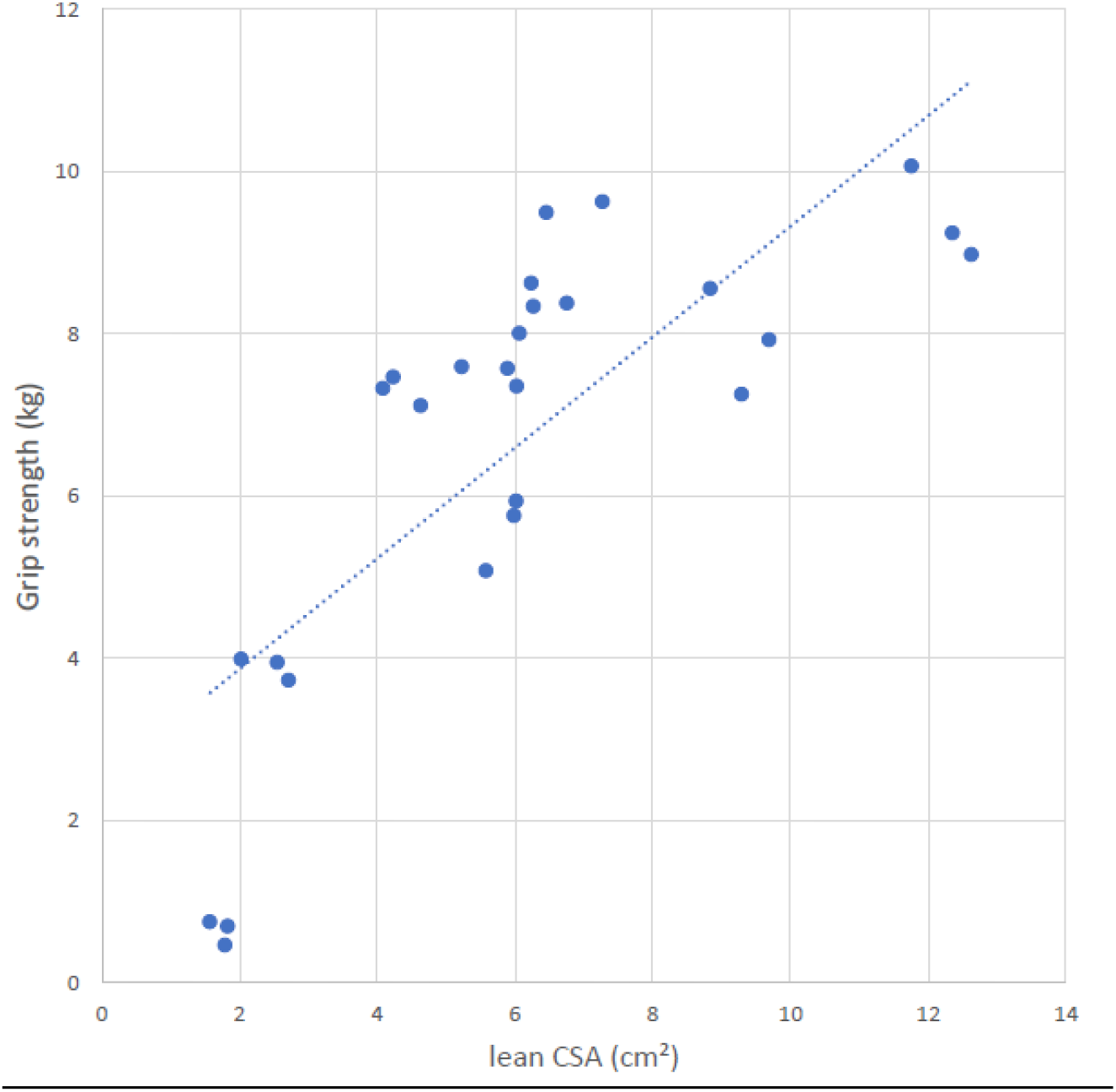
Correlation of the Grip Stength scores and the MRI data of the lean muscle area (non-fat) over the 24 week ATL1102 treatment period: The MRI is the determined lean muscle mass compartment across the mid (central) dominant forearm

## References

1. Crisafulli S, Sultana J, Fontana A, Salvo F, Messina S, Trifiro G. Global epidemiology of Duchenne muscular dystrophy: an updated systematic review and meta-analysis. Orphanet J Rare Dis. 2020;15(1):141.

2. Landfeldt E, Thompson R, Sejersen T, McMillan HJ, Kirschner J, Lochmuller H. Life expectancy at birth in Duchenne muscular dystrophy: a systematic review and meta-analysis. Eur J Epidemiol. 2020;35(7):643–53.

3. Bello L, Gordish-Dressman H, Morgenroth LP, Henricson EK, Duong T, Hoffman EP, et al. Prednisone/prednisolone and deflazacort regimens in the CINRG Duchenne Natural History Study. Neurology. 2015;85(12):1048–55.

4. Bello L, Kesari A, Gordish-Dressman H, Cnaan A, Morgenroth LP, Punetha J, et al. Genetic modifiers of ambulation in the Cooperative International Neuromuscular Research Group Duchenne Natural History Study. Ann Neurol. 2015;77(4):684–96.

5. Birnkrant DJ, Bushby K, Bann CM, Apkon SD, Blackwell A, Brumbaugh D, et al. Diagnosis and management of Duchenne muscular dystrophy, part 1: diagnosis, and neuromuscular, rehabilitation, endocrine, and gastrointestinal and nutritional management. Lancet Neurol. 2018;17(3):251–67.

6. Rosenberg AS, Puig M, Nagaraju K, Hoffman EP, Villalta SA, Rao VA, et al. Immune-mediated pathology in Duchenne muscular dystrophy. Sci Transl Med. 2015;7(299):299rv4.

7. Zanotti S, Gibertini S, Di Blasi C, Cappelletti C, Bernasconi P, Mantegazza R, et al. Osteopontin is highly expressed in severely dystrophic muscle and seems to play a role in muscle regeneration and fibrosis. Histopathology. 2011;59(6):1215–28.

8. Pinto-Mariz F, Carvalho LR, de Mello W, Araujo Ade Q, Ribeiro MG, Cunha Mdo C, et al. Differential integrin expression by T lymphocytes: potential role in DMD muscle damage. J Neuroimmunol. 2010;223(1-2):128–30.

9. Pinto-Mariz F, Rodrigues Carvalho L, Prufer De Queiroz Campos Araujo A, De Mello W, Goncalves Ribeiro M, Cunha Mdo C, et al. CD49d is a disease progression biomarker and a potential target for immunotherapy in Duchenne muscular dystrophy. Skelet Muscle. 2015;5:45.

10. Limmroth V, Barkhof F, Desem N, Diamond MP, Tachas G, Group ATLS. CD49d antisense drug ATL1102 reduces disease activity in patients with relapsing-remitting MS. Neurology. 2014;83(20):1780–8.

11. Lagrota-Candido J, Canella I, Savino W, Quirico-Santos T. Expression of extracellular matrix ligands and receptors in the muscular tissue and draining lymph nodes of mdx dystrophic mice. Clin Immunol. 1999;93(2):143–51.

12. Hogarth MW, Houweling PJ, Thomas KC, Gordish-Dressman H, Bello L, Cooperative International Neuromuscular Research G, et al. Evidence for ACTN3 as a genetic modifier of Duchenne muscular dystrophy. Nat Commun. 2017;8:14143.

13. Seferian AM, Moraux A, Annoussamy M, Canal A, Decostre V, Diebate O, et al. Upper limb strength and function changes during a one-year follow-up in non-ambulant patients with Duchenne Muscular Dystrophy: an observational multicenter trial. Plos One. 2015;10(2):e0113999.

14. Servais L, Deconinck N, Moraux A, Benali M, Canal A, Van Parys F, et al. Innovative methods to assess upper limb strength and function in non-ambulant Duchenne patients. Neuromuscul Disord. 2013;23(2):139–48.

15. Hogrel JY, Wary C, Moraux A, Azzabou N, Decostre V, Ollivier G, et al. Longitudinal functional and NMR assessment of upper limbs in Duchenne muscular dystrophy. Neurology. 2016;86(11):1022–30.

16. Mercuri E, Pichiecchio A, Counsell S, Allsop J, Cini C, Jungbluth H, et al. A short protocol for muscle MRI in children with muscular dystrophies. Eur J Paediatr Neurol. 2002;6(6):305–7.

17. Mercuri E, Pichiecchio A, Allsop J, Messina S, Pane M, Muntoni F. Muscle MRI in inherited neuromuscular disorders: past, present, and future. J Magn Reson Imaging. 2007;25(2):433–40.

18. Fischer D, Kley RA, Strach K, Meyer C, Sommer T, Eger K, et al. Distinct muscle imaging patterns in myofibrillar myopathies. Neurology. 2008;71(10):758–65.

19. Ricotti V, Evans MR, Sinclair CD, Butler JW, Ridout DA, Hogrel JY, et al. Upper Limb Evaluation in Duchenne Muscular Dystrophy: Fat-Water Quantification by MRI, Muscle Force and Function Define Endpoints for Clinical Trials. Plos One. 2016;11(9):e0162542.

20. Pane M, Coratti G, Brogna C, Mazzone ES, Mayhew A, Fanelli L, et al. Upper limb function in Duchenne muscular dystrophy: 24 month longitudinal data. Plos One. 2018;13(6):e0199223.

21. Tachas G, Desem N, Button P, Pane E, and Mercuri E, World Muscle Society 2020, P.284 ATL1102 treatment improves PUL2.0 in non-ambulant boys with Duchenne muscular dystrophy compared to a natural history control. Neuromuscular Disorders Volume 30, Supplement 1, S129-S130 October 1, 2020, http://dx.doi.org/10.1016/j.nmd.2020.08.28

