## Supplementary Fig. 4 for "A Phase 2 open-label study to determine the safety and efficacy of weekly dosing of ATL1102 in patients with non-ambulatory Duchenne muscular dystrophy"

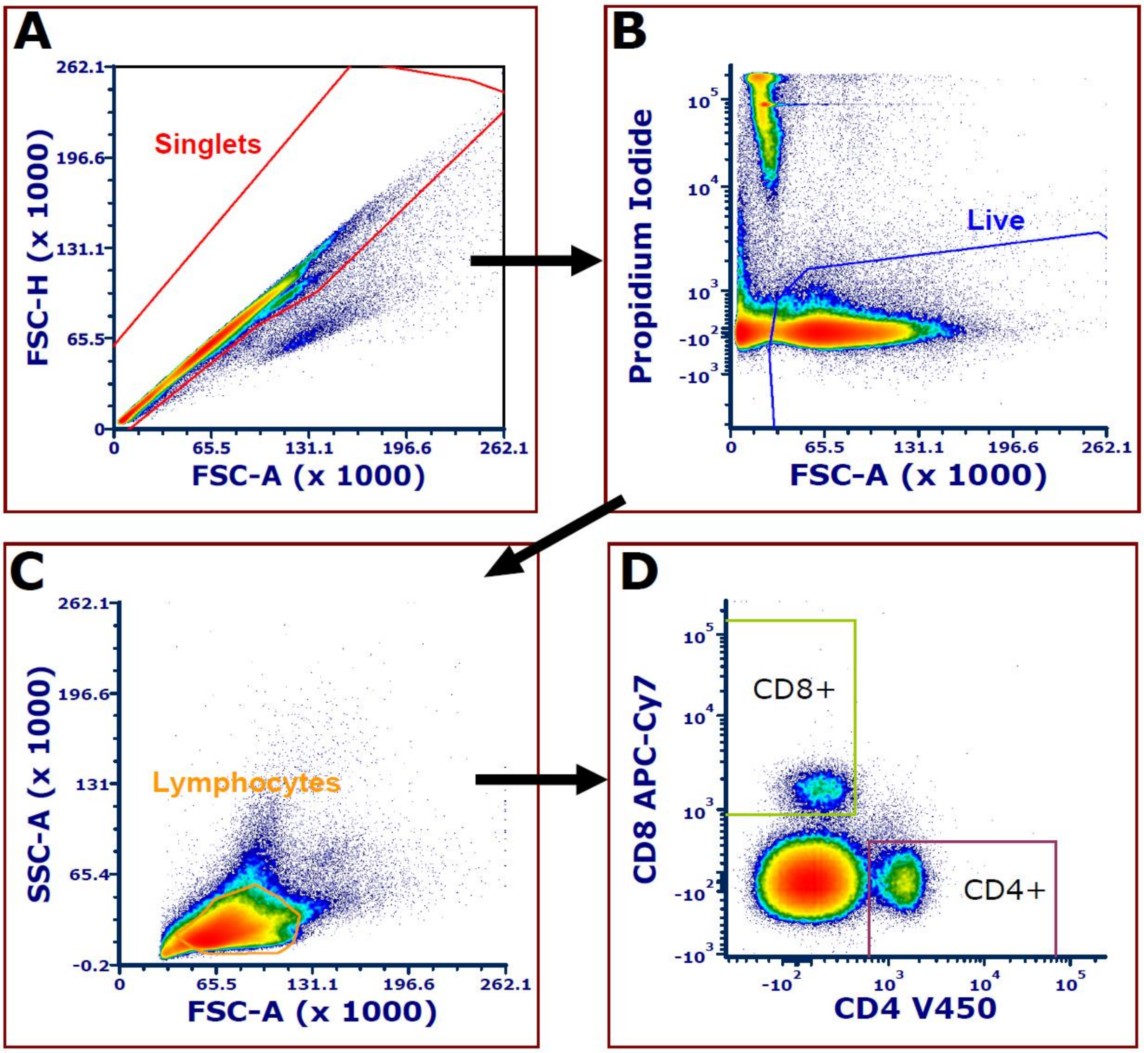


**Supplementary Fig. 4** - Gating strategy for identification of murine CD4+ and CD8+ T cell populations isolated from spleen of *mdx* mice treated with ISIS 348574. A) Singlets (FSC-H vs FSC-A), B) Live cells (Propidium Iodide vs FSC-A) and C) Lymphocytes (SSC-A vs FSC-A) were gated to remove doublets, dead cells, debris and large/granular cells. D) Anti-CD4-V450 and anti-CD8a APC-Cy7 to were used to gate populations of CD4+ and CD8+ T cells.
